# Assessment and staging of A/T/N with a single dynamic [^18^F]PI-2620 recording

**DOI:** 10.1101/2025.01.14.25320240

**Authors:** Johannes Gnörich, Julia Kusche-Palenga, Agnes Kling, Amir Dehsarvi, Angela Bronte, Lukas Frontzkowski, Artem Zatcepin, Mirlind Zaganjori, Florian Schöberl, Sebastian N Roemer, Boris-Stephan Rauchmann, Carolin Kurz, Carla Palleis, Alexander M Bernhardt, Alexander Jäck, Sabrina Katzdobler, Maximilian Scheifele, Theresa Bauer, Gérard N Bischof, Thilo van Eimeren, Alexander Drzezga, Jan Häckert, Robert Perneczky, Michael Rullmann, Katharina Bürger, Andreas Zwergal, Johannes Levin, Peter Bartenstein, Osama Sabri, Henryk Barthel, Sophia Stöcklein, Günter Höglinger, Nicolai Franzmeier, Matthias Brendel

## Abstract

Patients with Alzheimer’s disease (AD) and clinically overlapping neurodegenerative diseases are classified molecularly using the A/T/N classification system. Apart from fluid biomarkers and structural MRI, the three-dimensional A/T/N system incorporates characteristic features from β-amyloid-PET (A), tau-PET (T), and FDG-PET (N). We evaluated if dynamic features of tau-PET with [^18^F]PI-2620 allow assessment of A/T/N in individual patients using a single imaging session. Cortical tissue clearance (K2a) of [^18^F]PI-2620 was validated as a surrogate of the β-amyloid status against β-amyloid-PET and cerebrospinal fluid (CSF) Aβ_42/40_ ratio, demonstrating remarkable positive (91.5%) and negative (95.1%) predictive values at an AUC of 0.99 (P<0.0001). K2a outperformed cortical tau burden as a surrogate for β-amyloid status in 47 participants with a clinical diagnosis of probable AD (3/4-repeat(R)-tauopathy) and 82 β-amyloid-negative patients with primary 4R-tauopathies. Perfusion-like [^18^F]PI-2620 images (R1) were validated as a surrogate marker for neuronal injury, exhibiting strong quantitative and visual correlations with FDG-PET and early-phase β-amyloid-PET, as well as with volumetric MRI and CSF total tau levels. Composite quantitative A/T/N indices facilitated personalized staging along temporal disease trajectories. Our results suggest that [^18^F]PI-2620 imaging has the potential to facilitate the assessment of region and stage dependent PET-based A/T/N during a single dynamic PET session.

**Graphical Abstract:** 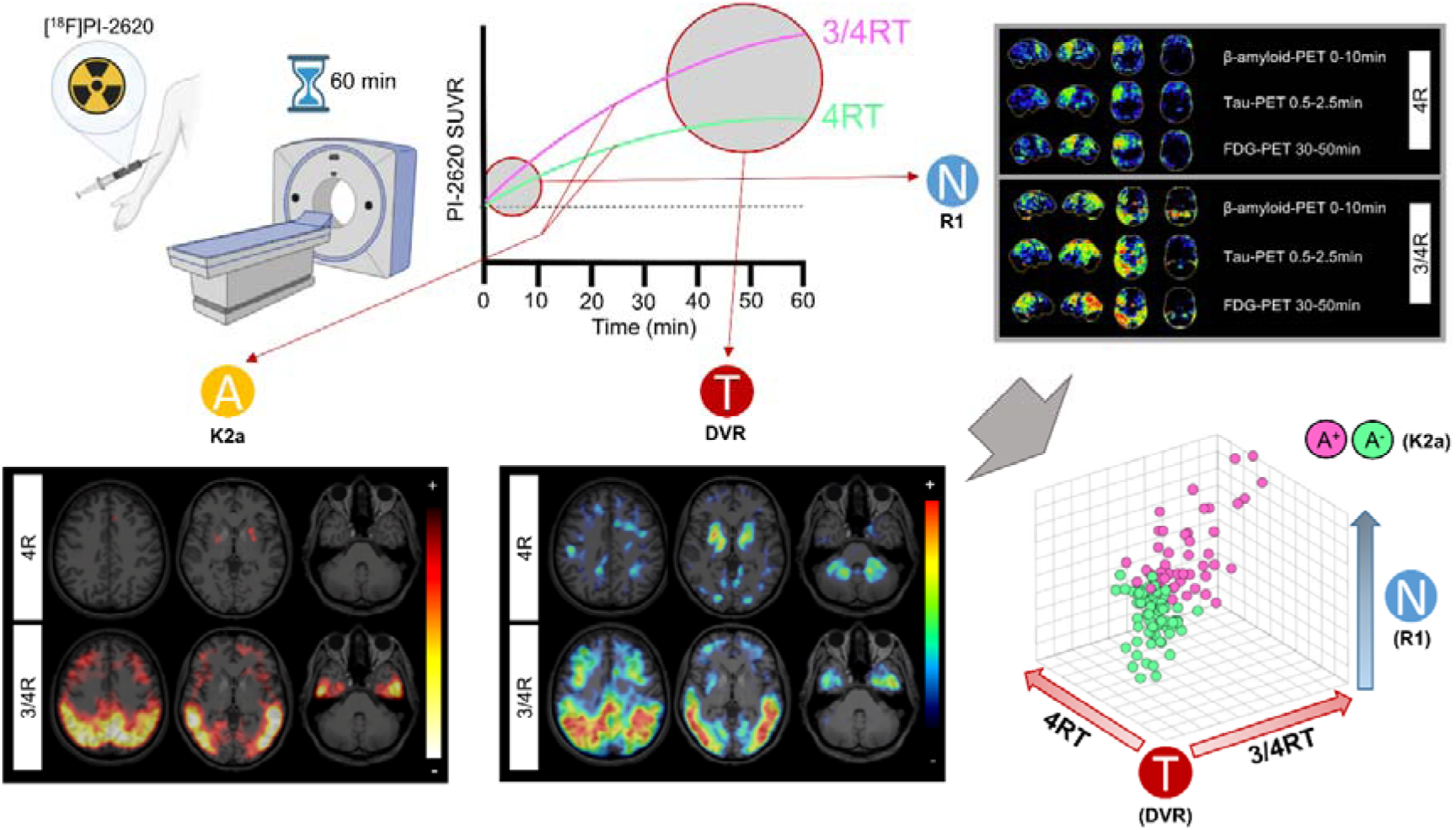

## Introduction

Tauopathies comprise a spectrum of neurodegenerative diseases characterized by the aggregation of tau protein in the brain. These diseases are classified into primary and secondary tauopathies based on the molecular composition of tau aggregates and associated upstream protein deposits. Primary tauopathies, such as progressive supranuclear palsy (PSP) and corticobasal degeneration (CBD), are characterized by the abnormal aggregation of tau isoforms with 4-repeat (4R) binding domains, leading to distinct clinical phenotypes including atypical parkinsonian syndromes and cognitive impairments [1–3]. On the other hand, Alzheimer’s disease (AD) represents a secondary tauopathy characterized by the upstream co-occurrence of beta amyloid plaque pathology followed by the intraneuronal aggregation of 3/4R-tau primarily in the cortex, resulting in neurodegeneration and cognitive decline [4, 5]. However, imaging and fluid biomarkers are less advanced for in vivo classification of primary 4R-tauopathies (4RT), limiting our mechanistic understanding and impeding treatment development.

The emergence of the second-generation tau-PET tracer [^18^F]PI-2620 holds significant promise for in vivo detection of tauopathies. Unlike first-generation tau-PET ligands, [^18^F]PI-2620 exhibits reduced off-target binding, making it a more specific tool for detecting tau pathology [6]. Its’ high affinity for both 3R and 4R aggregated tau isoforms, as demonstrated in vitro and in vivo studies, positions it as a valuable biomarker capable of discerning between various tauopathies [7–9]. While AD-tau filaments exhibit favourable energetic and kinetic properties for [^18^F]PI-2620 binding, interactions with CBD- and PSP-tau filaments are kinetically weaker [10]. This was also demonstrated by dynamic PET parameters derived from non-invasive reference tissue modelling, which revealed faster tracer clearance in 4R-tauopathies, suggesting less stable binding compared to 3/4R-tauopathies. Notably, the parameter K2a, representing tracer efflux from the tissue, indicated potential in differentiating between distinct tauopathies, serving as an indirect marker for β-amyloid pathology prediction, enhancing the accuracy of differential diagnoses [11]. As a direct result, earlier imaging times (i.e., 20-40 min p.i.) provide higher quantitative and visual sensitivity for detection of patients with PSP and CBD, compared to later imaging times [12]. Furthermore, [^18^F]PI-2620 holds promise as a surrogate biomarker for neuronal injury in tauopathies, as evidenced by its ability to demonstrate decreased early-phase [^18^F]PI-2620 perfusion [13, 14]. The integration of perfusion and tau biomarkers enhanced biomarker-guided stratification of tauopathies within diagnostic algorithms and demonstrated improved discrimination, though it has yet to be prospectively validated [9].

These recent advancements in biomarker development, particularly in AD, have revolutionized disease diagnosis and monitoring, facilitating the establishment of diagnostic workflows and biomarker-based staging systems, as initially proposed by Jack et al. [15]. The A/T/N classification scheme offers a comprehensive framework for characterizing the underlying pathology. This pivotal scheme stratifies neurodegenerative diseases based on the presence of β-amyloid (A), tau (T), and neurodegeneration (N) biomarkers, as measured by imaging techniques and fluid biomarker assessments. Imaging by β-amyloid-PET (A), tau-PET (T), and FDG-PET for (N) can provide not only binary classification, but also regional staging of A/T/N biomarker abnormality. However, this approach requires multiple imaging sessions, posing an added burden to patients and the healthcare system. For this reason, there is still an unmet need to develop a straightforward solution that enables a ‘one-stop-shop’ approach for classifying patients based on A/T/N criteria.

In this study, we assessed the value of dynamic [^18^F]PI-2620 tau-PET imaging by utilization of the i) kinetics of radiotracer distribution as β-amyloid surrogate (A), ii) late phase binding to tau (T), and iii) perfusion phase as a neurodegeneration surrogate (N). Our approach extends beyond traditional AD frameworks by applying the A/T/N system also to 4RT, such as PSP and CBD. This focus on 4RT addresses a critical gap in personalized neurodegenerative diagnostics, as these disorders often present distinct biomarker profiles compared to AD. However, the differentiation between tauopathies can be challenging due to significant clinical and pathological overlap with atypical forms of AD, often exhibiting tau pathology patterns that can mimic those seen in primary tauopathies. This overlap complicates diagnosis, particularly in the early stages of disease progression, and may lead to misclassification without comprehensive biomarker assessment.

Thus, we evaluated staging by all three A/T/N biomarkers in individual patients within a single dynamic imaging session. We validated our results against gold standard assessments, which involved integrating multimodal PET imaging (β-amyloid & FDG), volumetric MRI, and cerebrospinal fluid (CSF) biomarkers (p-tau-181, t-tau & Aβ_42/40_). Finally, we developed a 3-dimensional quantitative staging index to determine personalized profiles of A/T/N-based disease severity.

## Results

### Demographics of the in vivo dynamic [^18^F]PI-2620 tau-PET imaging population

Patients included belong to the AD continuum (total n=47; ⌀75±9 years, 62% female) as 3/4RT or PSP/CBS (n=82; ⌀75±7 years, 43% female) as 4RT and were compared to healthy controls (HC) (n=17; ⌀70±10 years, 53% female). Demographics of the study cohort are reported in **Table 1**.

**Table 1.** Demographics at the group level. Abbreviations: 4RT = 4-repeat-tauopathy, 3/4RT = 3/4-repeat-tauopathy, CBS = corticobasal syndrome, CSF = cerebrospinal fluid, FDG = Fluorodeoxyglucose, NA = not applicable, MCI = mild cognitive impairment, MoCA = Montreal Cognitive Assessment, MRI = Magnetic resonance imaging, PET = positron emission tomography, PSP = progressive supranuclear palsy, PSP RS = PSP rating scale, p-tau-181 = phosphorylated tau, SD = standard deviation, t-tau = total tau.

| Demographic | 4RT | 3/4RT | Healthy controls |
| --- | --- | --- | --- |
| No. | 82 | 47 | 17 |
| Subgroups | PSP (n=71),<br>CBS (n=11) | MCI (n=14),<br>dementia (n=33) | NA |
| Age, mean (SD), y | 75 (7) | 75 (9) | 70 (10) |
| Sex, No. (%) |  |  |  |
| Female | 35 (43) | 29 (62) | 9 (53) |
| Male | 47 (57) | 18 (38) | 8 (47) |
| Dynamic [<sup>18</sup>F]PI-2620 PET | 82 | 47 | 17 |
| FDG-PET | 36 | 15 | NA |
| β-amyloid-PET | 47 | 31 | NA |
| Volumetric MRI | 43 | 22 | NA |
| CSF Aβ<sub>42/40</sub> | 63 | 41 | NA |
| CSF p-tau-181 & t-tau | 56 | 39 | NA |
| MoCA score, mean (SD) | 21.7 (4.8) | 16.7 (6.2) | NA |
| PSP RS score, mean (SD) | 30.6 (12.6) | 24.6 (7.4) | NA |

### K2a images show distinct and specific patterns in tauopathies

First, we compared clearance characteristics of [^18^F]PI-2620 based on K2a, which was successfully derived for all individuals using kinetic modelling [9, 16]. Visual assessment revealed similar K2a maps of 4RT compared to HC with slightly lower tracer clearance in the basal ganglia (**Figure 1A-B**). In contrast, 3/4RT (i.e. AD patients) exhibited significantly reduced tracer clearance in cortical regions suggesting strong and irreversible tracer binding to AD-type tau, which demonstrated high sensitivity even in individuals with limited late-phase tau-PET signals. In individual β-amyloid-negative patients with PSP-CBS and CBS, no relevant reduction of tracer efflux was observed, even if the basal ganglia exceeded DVR z-scores of 2 (**Figure 2A**). In contrast, individual β-amyloid-positive patients with AD-CBS demonstrated elevated DVR in cortical regions. Here, the K2a signal even extended beyond the area of DVR elevation, being widely detectable in distant cortical brain regions (**Figure 2A**).

**Figure 1:**
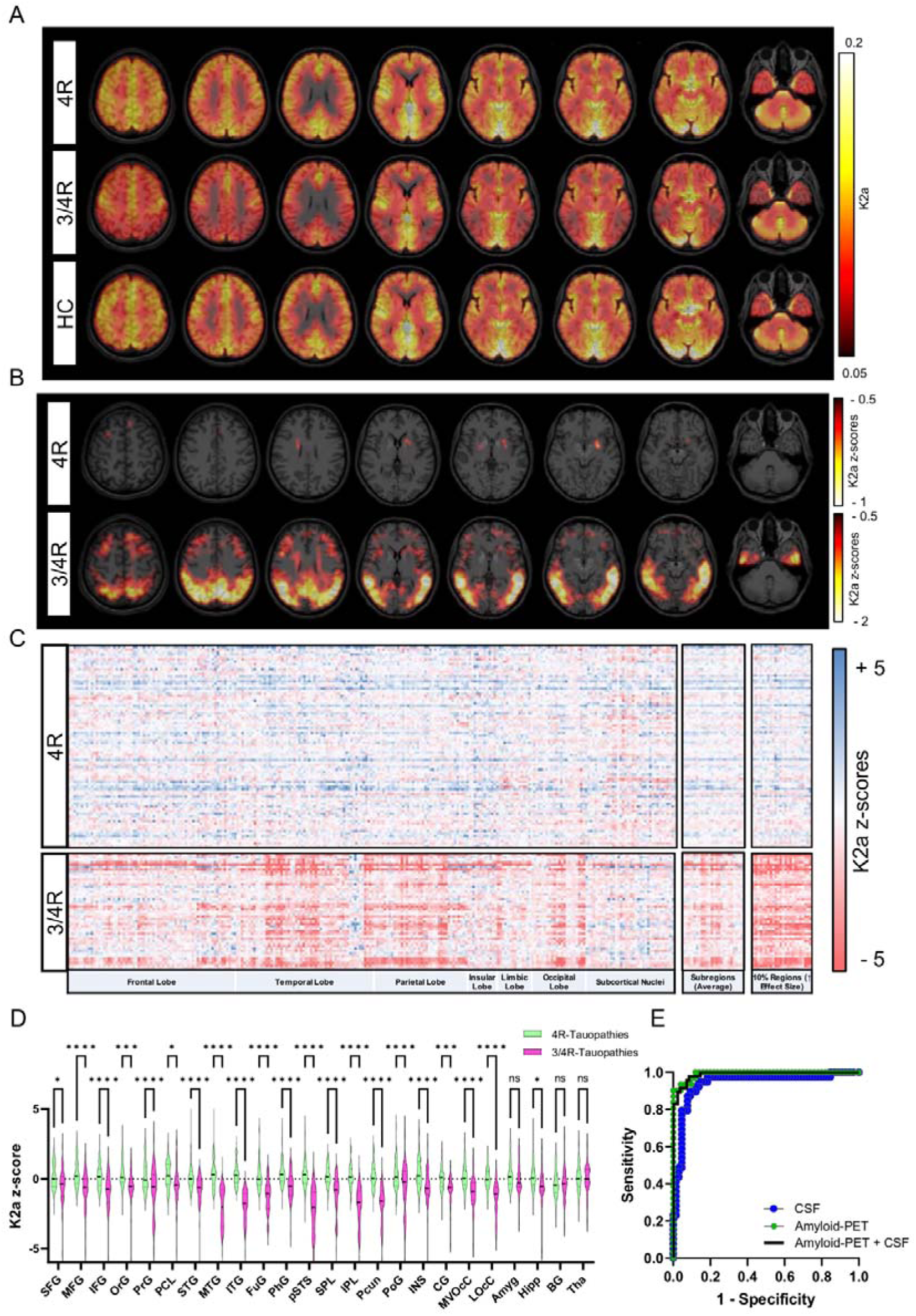
K2a assessment as an index of tracer clearance in tauopathies. (**A-B**) Average [^18^F]PI-2620 K2a and K2a z-score images of 4RT (n=82), 3/4RT (n=47) in relation to healthy controls (n=17) (HC), illustrated by axial slice overlays on a standard magnetic resonance imaging (MRI) template in MNI space. (**C**) Heatmaps on the left show K2a z-scores of all Brainnetome atlas regions among all individuals. Heatmaps on the right demonstrate the upper 10% of discriminating regions determined by effect size analysis between both tauopathy cohorts. Intermediate heatmaps illustrate the composite Brainnetome atlas subregions, which are subsequently shown in the ANOVA analysis of panel (**D**). (**E**) ROC curves represent the results of a logistic regression analysis using the averaged K2a of the top 10% discriminating regions against CSF (blue dots), β-amyloid-PET (green dots), and both combined (black line) to predict β-amyloid positivity in all participants. Abbreviations: SFG = Superior Frontal Gyrus, MFG = Middle Frontal Gyrus, IFG = Inferior Frontal Gyrus, OrG = Orbital Gyrus, PrG = Precentral Gyrus, PCL = Paracentral Lobule, STG = Superior Temporal Gyrus, MTG = Middle Temporal Gyrus, ITG = Inferior Temporal Gyrus, FuG = Fusiform Gyrus, PhG = Parahippocampal Gyrus, pSTS = Posterior Superior Temporal Sulcus, SPL = Superior Parietal Lobule, IPL = Inferior Parietal Lobule, Pcun = Precuneus, PoG = Postcentral Gyrus, INS = Insular Gyrus, CG = Cingulate Gyrus, MVOcC = MedioVentral Occipital Cortex, LOcC = Lateral Occipital Cortex, Amyg = Amygdala, Hipp = Hippocampus, BG = Basal Ganglia, Tha = Thalamus.

**Figure 2:**
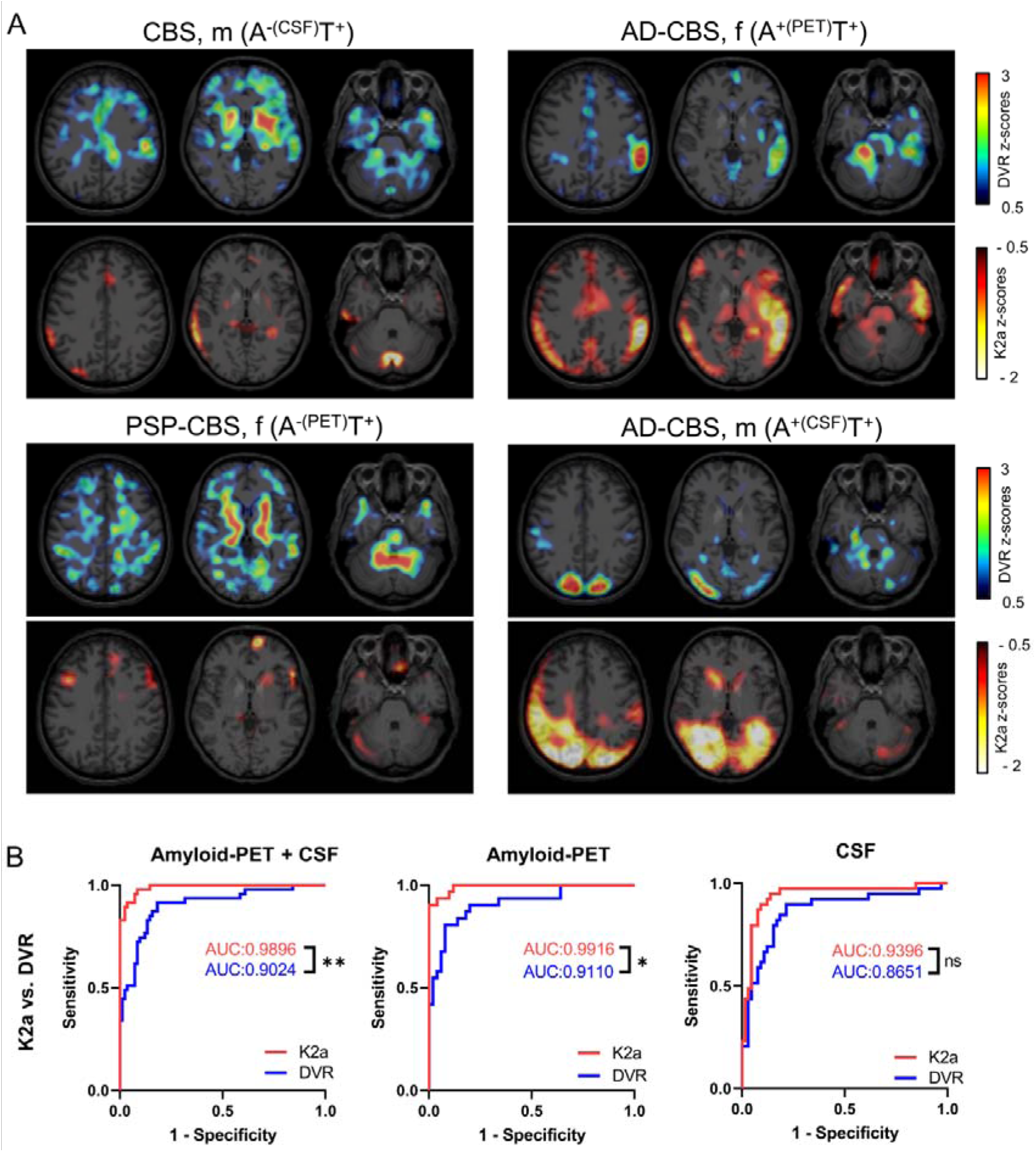
Tau-PET tracer clearance for the prediction of β-amyloid positivity. (**A**) Four sex-and age-matched individuals (range 75-82 years) with clinically probable tauopathies and cortical tau-PET binding were distinguished according to their K2a patterns. Interestingly, off-target binding spots found in the vermal region of the β-amyloid-positive cases did not show significantly altered tracer clearance. (**B**) Comparison of ROC curves between K2a (red line) and DVR (blue line) for the top 10% discriminating regions from the Brainnatome atlas in predicting β-amyloid positivity across all participants, as assessed by gold standard methods.

### [^18^F]PI-2620 tracer clearance predicts β-amyloid positivity in tauopathies (A)

All K2a z-score values derived from the Brainnetome atlas (n=246), summarized anatomical subregions (n=24), and the 10% most discriminating ROIs (effect size analysis between 4RT and 3/4RT, n=24) are visualized for all individual patients in **Figure 1C**. Significant differences of [^18^F]PI-2620 clearance among the two study cohorts were observed in 21 out of 24 subregions. Effect size analysis between both cohorts (4RT versus 3/4RT) revealed highest discriminative power in cortical regions, best represented by the inferior temporal gyrus (Cohen’s d=2.49) (**Figure 1C-D**). 18 out of 24 (75%) of the 10% most discriminative ROIs were identical to the 10% ROIs with the lowest tracer clearance in 3/4RT.

Next, we questioned if quantitative [^18^F]PI-2620 tracer clearance is able to predict the β-amyloid status as assessed by β-amyloid-PET scans (n=81) and/or CSF Aβ_42/40_ ratio <5.5 (n=104). In cases with both, Aβ-CSF and Aβ-PET available, we observed a congruency of 86% (32/37) in 3/4RT and 97% (33/34) in 4RT, when PET was used as the decisive read-out [17]. Averaged K2a values of the 24 pre-assessed discriminative ROIs were able to predict the β-amyloid status at a positive predictive value (PPV) of 91.5% and a negative predictive value (NPV) of 95.1% at an AUC of 0.99 (P<0.0001, **Figure 1E**). In a sub-analysis of patients with β-amyloid-PET, stronger PPV (93.6%), NPV (96.0%), and AUC (0.99, P<0.0001) were observed compared to a CSF sub-analysis (PPV: 88.6%, NPV 88.4%, AUC: 0.94, P<0.0001, **Figure 1E**).

To assess whether K2a predicts β-amyloid positivity more effectively than tau binding alone, we compared the AUC of K2a to DVR, demonstrating that K2a provided stronger discrimination of β-amyloid positivity in relation to β-amyloid-PET (P=0.013), Aβ-CSF (P=0.10) and both combined assessments (P=0.0033) (**Figure 2B**).

### [^18^F]PI-2620 binding indicates distinct topology among different tauopathies (T)

As expected from our previous research [9], significant differences in [^18^F]PI-2620 binding among both study cohorts, utilizing DVR, were observed in 11/24 Brainnetome subregions, all located within the cortex. Interestingly, the regional effect size analysis again revealed the highest difference among all ROIs in the inferior temporal gyrus (Cohen’s d=1.85, compared to d=2.49 for K2a) (**Figure 3A-C**). Again, a ROC analysis was performed, now using the averaged DVR of the top 10% of ROIs with the highest discriminatory power to test whether [^18^F]PI-2620_DVR_ successfully predict β-amyloid positivity. AUC of DVR β-amyloid status prediction was 0.91 (P<0.0001) for validation against β-amyloid-PET, 0.87 (P<0.0001) for CSF and 0.90 (P<0.0001) for both modalities combined (**Figure 3D**).

**Figure 3:**
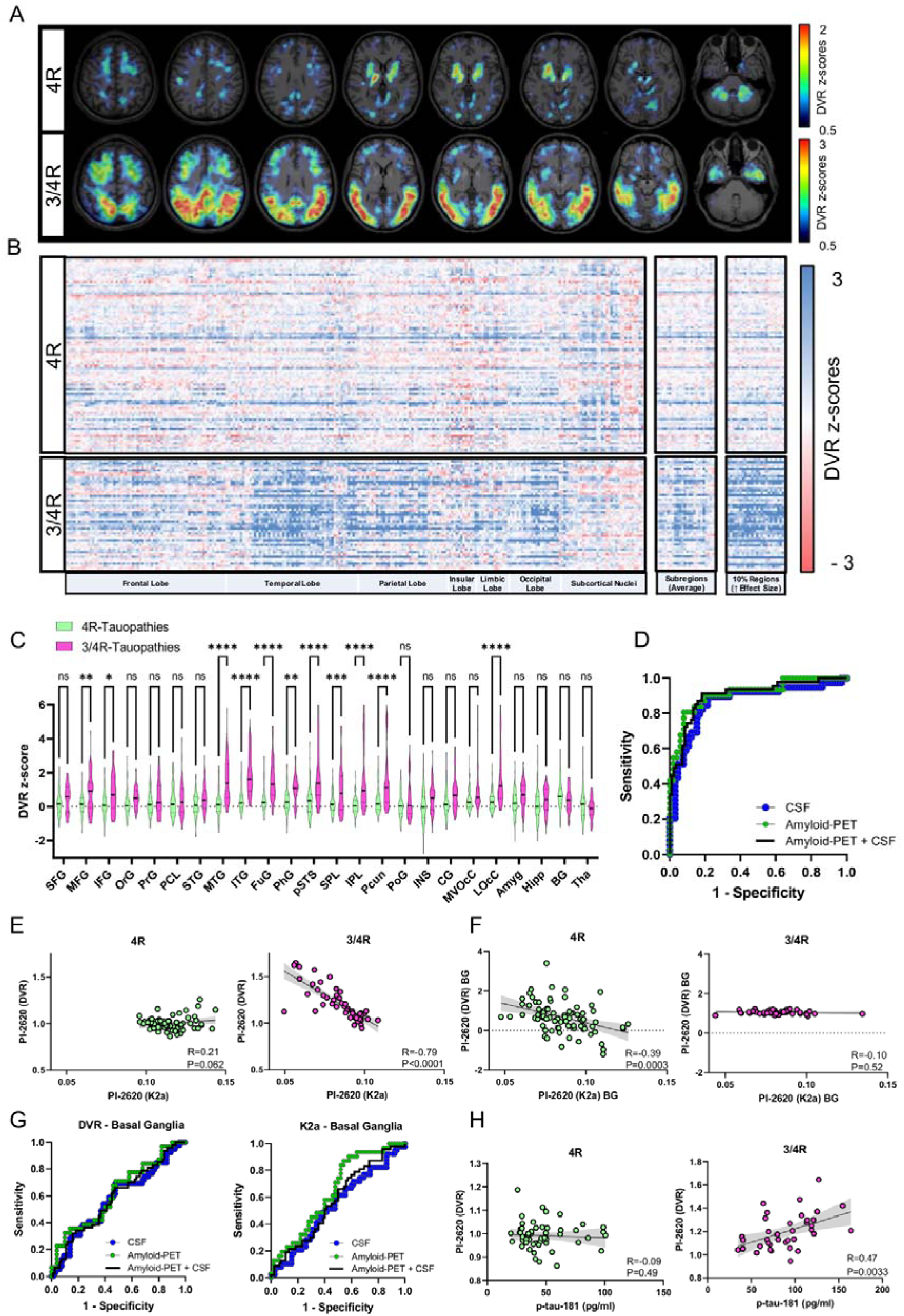
[^18^F]PI-2620 binding for the detection of tau accumulation patterns in 4RT and 3/4RT. (**A**) Average [^18^F]PI-2620 DVR z-score maps of 4RT and 3/4RT, illustrated by axial slice overlays on a standard MRI template in MNI space. (**B**) Heatmaps on the left show DVR z-scores of all Brainnetome atlas regions for all individuals. Heatmaps on the right show the top 10% of discriminating regions determined by effect size analysis between both cohorts. Intermediate heatmaps illustrate the composite Brainnetome atlas subregions. (**C**) ANOVA analysis of Brainnetome atlas subregions. (**D**) ROC curves represent the results of a logistic regression analysis using DVR of the top 10% discriminating regions against CSF (blue dots), β-amyloid-PET (green dots), and both combined (black line) to predict β-amyloid positivity in all patients. (**E-F**) Correlation plots visualize association between [^18^F]PI-2620 K2a and DVR in the 10% most discriminating K2a-based regions and the basal ganglia solely. (**G**) ROC analysis of [^18^F]PI-2620 K2a and DVR derived from the basal ganglia, validated against amyloid gold standard assessments. (**H**) Scatterplots depict association between [^18^F]PI-2620 DVR and p-tau-181 from CSF in both study cohorts.

To deeper understand the proposed biomarker of tracer clearance, we explored the association between [^18^F]PI-2620 tau binding and K2a in the predefined K2a-based discriminatory ROIs. While DVR and K2a only showed a trend towards correlation in patients with 4RT (R=0.21; P=0.062), a strong negative association between tau binding and tracer clearance was observed in patients with 3/4RT (R=-0.79; P<0.0001) (**Figure 3E**). In the basal ganglia, K2a was negatively associated with DVR in patients with 4RT (R=-0.39; P=0.0003), but not in patients with 3/4RT (R=-0.10; P=0.52) (**Figure 3F**). ROC analysis of K2a in the basal ganglia revealed significant prediction of β-amyloid PET positivity as a gold standard (AUC=0.64; P=0.030), but not when β-amyloid in CSF (AUC=0.54; P=0.51) or both assessments (AUC=0.57; P=0.17) were considered for determination of the β-amyloid status. Similarly, [^18^F]PI-2620 DVR in the basal ganglia did not show significant prediction of the β-amyloid status in PET (AUC=0.62; P=0.07), CSF (AUC=0.58; P=0.19), or when assessed by both biomarkers (AUC=0.58; P=0.11) (**Figure 3G**). There was no significant correlation between DVR and p-tau-181 in the discriminatory regions of 4RT (R=-0.09; P=0.49), but a significant correlation in the 3/4RT group (R=0.47; P=0.0033) (**Figure 3H**), along with the overall elevated p-tau-181 levels in 3/4RT compared to the 4RT group (P<0.0001) (**Supplemental Figure 4**).

### Delivery of [^18^F]PI-2620 for detection and staging of neuronal injury in tauopathies (N)

Early-phase [^18^F]PI-2620 has shown promising results in identifying neuronal injury in patients with tauopathies, demonstrating correlations with established indices of neurodegeneration in small mixed samples [13] and providing intriguing insights in 4RT [14]. Additionally, early-phase tau-PET provides similar regional information compared to early-phase β-amyloid-PET [18], further supporting its role in diagnostic algorithms [9]. However, despite these encouraging findings, the application of [^18^F]PI-2620 has not yet been systematically validated in larger cohorts and in conjunction with β-amyloid and tau assessment. Thus, we aimed to comprehensively evaluate the utility of [^18^F]PI-2620 in this A/T/N evaluation of patients with tauopathies. Specifically, we assessed the dynamic parameter of tracer delivery, relative perfusion (R1), as a biomarker for neuronal damage, expanding our understanding of its diagnostic potential (**Supplemental Results, Supplemental Figures 1-3**).

To assess the potential of tracer delivery in staging neuronal damage, we examined tracer perfusion within a combined global mask including AD (CenTauR) [19] and PSP (Kovacs) [20, 21] target regions. This assessment was performed alongside with staging of the individual patient’s tau burden, as obtained by evaluation of tau-PET DVR for each of the two distinct sets of composite regions. After categorizing participants as β-amyloid-positive or negative according to the K2a cutoff determined by Youden’s index, a 3D scatter plot was generated, which effectively visually distinguished AD cases from primary tauopathies. The analysis of exemplary cases further illustrated these findings, with tau-PET binding patterns in cortical regions contributing to a higher CenTauR score, while enhanced uptake in the basal ganglia was associated with a higher Kovacs score (**Figure 4**). The results of the entire cohort suggested that 3-dimensional quantitative staging can determine personalized trajectories of A/T/N-based disease severity (**Figure 4**).

**Figure 4:**
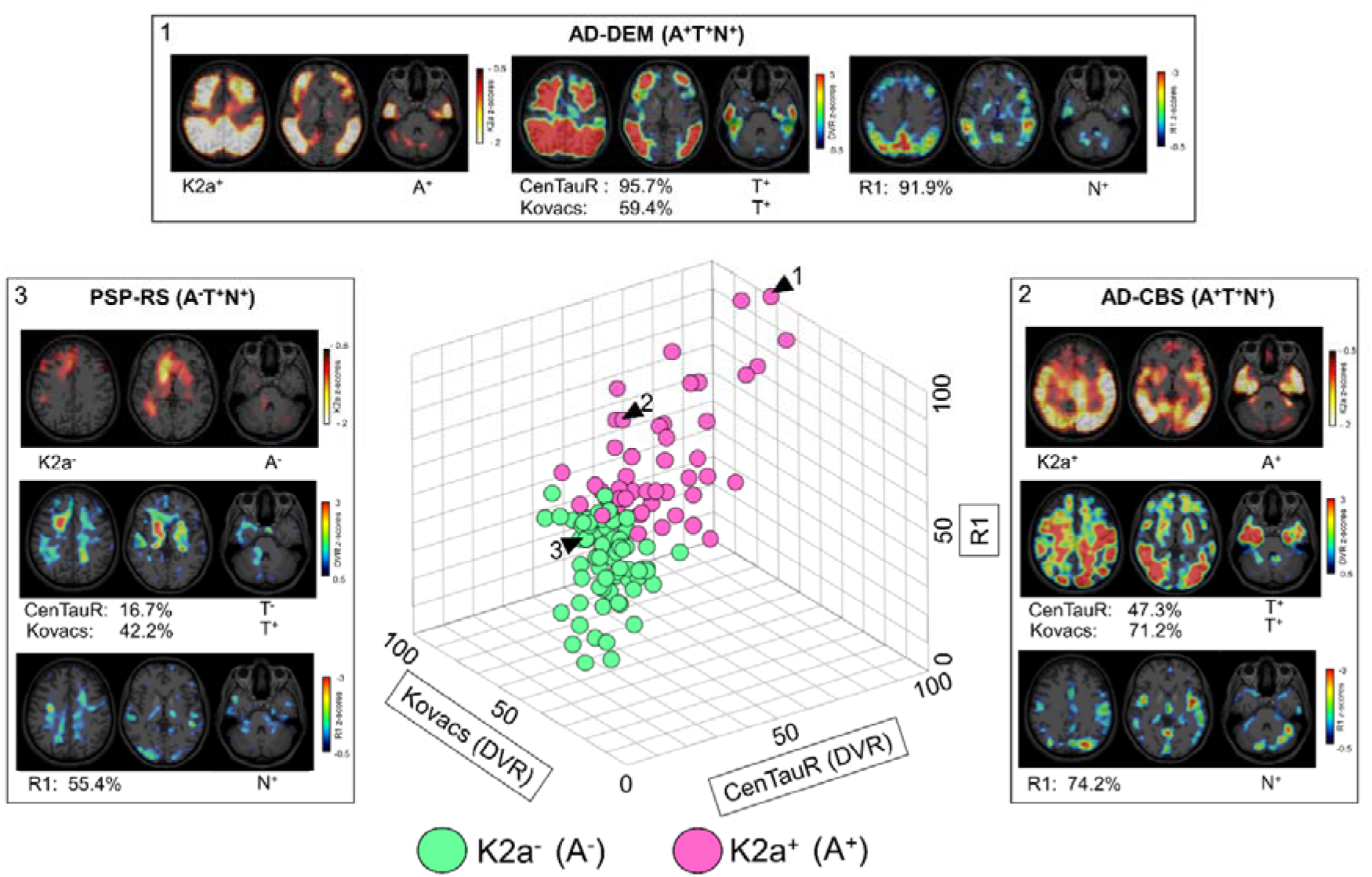
3-dimensional quantitative staging determines personalized trajectories of A/T/N-based disease severity in patients with tauopathies. Axial slices of representative patients with tauopathies with varying tau burden in the CenTauR and Kovacs masks are shown alongside K2a and R1 z-score images. The percentage values are calculated based on scaling relative to the individual with the highest and lowest values. 3D scatter plot visualizes all n=129 participants, with K2a predicted β-amyloid-positivity (purple dots) and β-amyloid-negativity (green dots).

A subset of both cohorts received additional FDG-PET_30-50min_ (4RT, n=36; 3/4RT, n=15), β-amyloid-PET_0-10min_ (4RT, n=37; 3/4RT, n=26) and CSF analysis with total-tau (t-tau) examination (4RT, n=56; 3/4RT, n=39), which served for gold standard validation of early-phase [^18^F]PI-2620 assessment as a surrogate of neurodegeneration. The intermodal correlation analysis revealed significant relationships across all major regions studied. The strongest correlations were observed between early-phase [^18^F]PI-2620 and FDG-PET (R=0.53, P<0.0001), as well as between early-phase [^18^F]PI-2620 and β-amyloid-PET_0-10min_ (R=0.56, P<0.0001) in the parietal cortex. (**Supplemental Results, Supplemental Figure 1**). Evaluation of t-tau indicated an elevation in the 3/4R cohort compared to the 4R (p<0.0001), along with a negative agreement with whole brain [^18^F]PI-2620_R1_ in 4R (R=-0.29; P=0.031) and 3/4R (R=-0.35; P=0.031) (**Supplemental Figure 2, 4**). In conclusion, quantitative staging of ATN with a single dynamic [^18^F]PI-2620 acquisition could enable detailed characterization of the individual patient’s biomarker trajectory.

### High visual agreement among early-phase [^18^F]PI-2620 and β-amyloid-PET as well as FDG-PET (N)

In terms of clinical translation of the proposed single session A/T/N paradigm, the assessment of tracer efflux will require computational tools, whereas tau-PET can be evaluated visually [21, 22]. Hence, we asked if neurodegeneration can also be obtained from early-phase tau-PET imaging by a visual read. Visual assessment was performed by three experienced readers evaluating global mean normalized 3D-SSP images of early-phase [^18^F]PI-2620_0.5–2.5min_, β-amyloid-PET_0–10min_ and FDG-PET (**Figure 5A)**. Representative images of early-phase [^18^F]PI-2620_0.5–2.5min_, β-amyloid-PET_0–10min_ and FDG-PET 3D-SSP are visualized in **Figure 5B**. First, we explored the tracer specific ratings among the readers using Fleiss kappa. The highest degree of congruency was attained for FDG-PET scans of 3/4RT (k=0.87; p<0.001), 4RT (k=0.83; p<0.001), and both cohorts combined (k=0.84; p<0.001). A high agreement was also found among the early-phase β-amyloid-PET reads of 3/4RT (k=0.75; p<0.001), 4RT (k=0.50; p<0.001), and both groups combined (k=0.59; p<0.001). Similarly, early-phase tau-PET reads indicated a strong congruency in 3/4RT (k=0.59; p<0.001), 4RT (k=0.60; p<0.001), and both groups (k=0.60; p<0.001). Next, we investigated the disease specific ratings among the different PET modalities using Cohen’s kappa. FDG-PET vs. early-phase β-amyloid-PET indicated the highest degree of agreement in the 3/4RT (k=1.0; p<0.001) followed by FDG-PET vs. early-phase tau-PET (k=0.59; p<0.001), and both early-phase reads (k=0.48; p<0.001). In patients with 4RT we found the strongest agreement between FDG-PET vs. early-phase tau-PET (k=0.74; p<0.001), followed by FDG-PET vs. early-phase β-amyloid-PET (k=0.67; p<0.001) and complemented by the couple of both early-phase PET modalities (k=0.54; p<0.001) (**Figure 5C**). In summary, visual assessment of early-phase tau-PET imaging delivers a reliable and robust surrogate of neurodegeneration across primary and secondary tauopathies.

**Figure 5:**
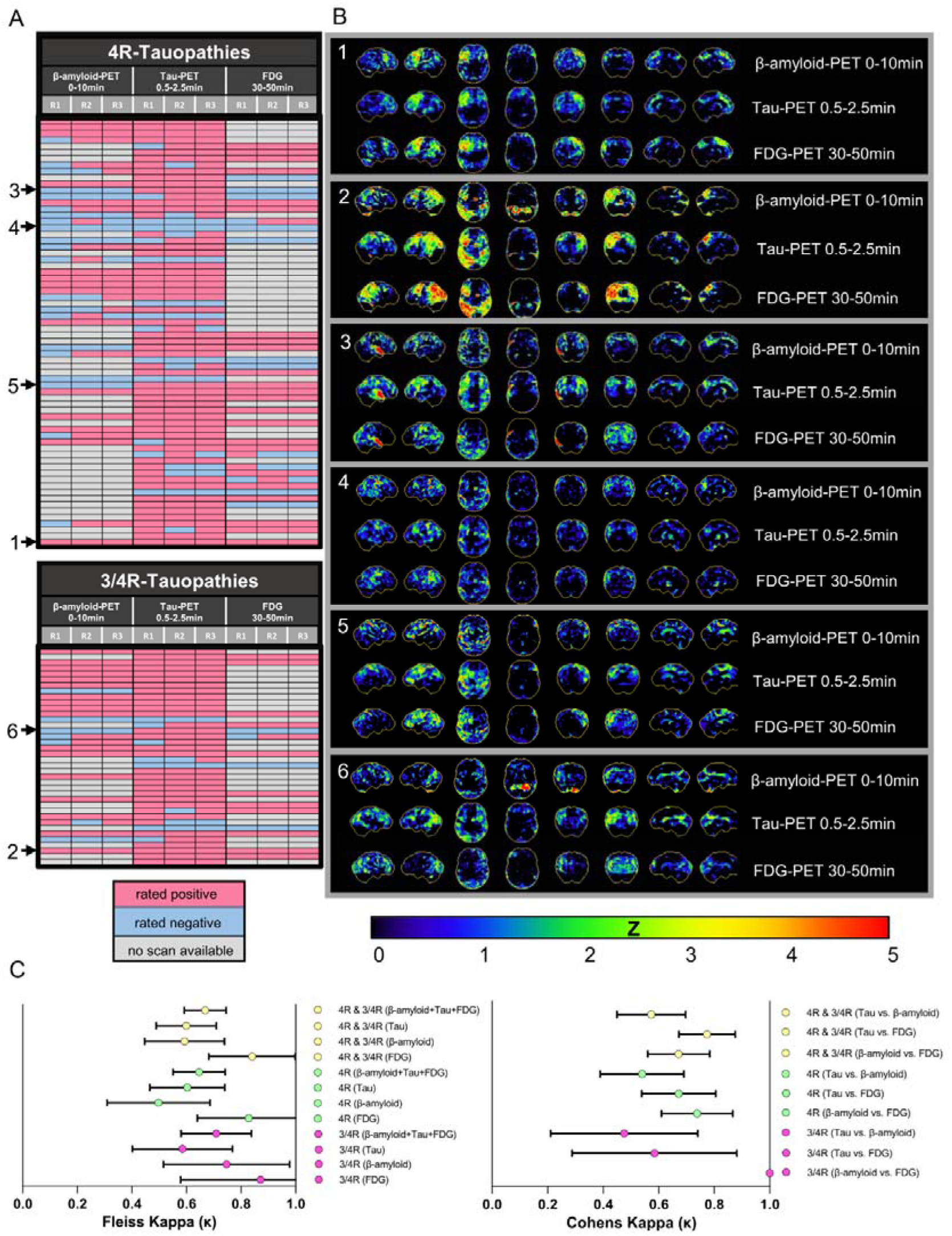
Visual evaluation of early-phase [^18^F]PI-2620 PET for detection of neurodegeneration. (**A**) Heatmaps of visual classification performed by dichotomous rating of [^18^F]PI-2620_0.5–2.5min_, β-amyloid-PET_0–10min_ and FDG-PET scans by three independent readers, who defined positivity (N+) or negativity (N-) for neuronal injury. (**B**) Representative 3D-SSP images (z-score maps) of congruent [^18^F]PI-2620_0.5–2.5min_, β-amyloid-PET_0–10min_ and FDG-PET scans. **B1**: 4RT rated *N+* by all three readers; **B2**: 3/4RT rated *N+* by all three readers; **B3**: 4RT rated *N+* by all three readers only in [^18^F]PI-2620_0.5–2.5min;_ PET; **B4**: 4RT rated *N-* by all three readers; **B5**: 4RT rated *N+* only in [^18^F]PI-2620_0.5–2.5min_ and FDG-PET; **B6**: 3/4RT rated *N+* by all three readers only in [^18^F]PI-2620_0.5–2.5min._ (**C**) Degree of agreement between the three readers for all three imaging modalities and both study cohorts depicted by Fleiss kappa (κ). Degree of congruency between two different scan modalities for both study cohorts was measured using Cohens Kappa (κ).

## Discussion

Molecular imaging is becoming increasingly prominent in definition schemes of neurodegenerative diseases, particularly AD [23] but also for biological classification of PD [24, 25]. The evolving diagnostic criteria for AD, particularly the revised framework proposed by Jack et al. [23], emphasize using a single Core 1 biomarker for diagnosis across the disease continuum. This biological-centric approach aligns with the broader A/T/N classification but raises questions about its clinical implications. Recent critiques, argue that focusing solely on β-amyloid (A) positivity may overlook the complexity of tau (T) and neurodegeneration (N) biomarkers, potentially leading to premature diagnoses in asymptomatic individuals [26, 27]. These concerns underscore the importance of integrating a comprehensive A/T/N assessment, rather than relying on a single biomarker, to capture the dynamic interplay of β-amyloid, tau, and neurodegenerative processes. Our findings support this holistic view, demonstrating that multi-faceted tau-PET metrics provide a robust framework for disease staging and progression monitoring, ensuring that diagnostic criteria reflect both biological and clinical realities. Therefore, it is essential to establish discriminative biomarkers that characterize the disease trajectory of the individual patient by extracting the maximum information from each PET examination to minimize cost and burden to individual patients and the healthcare system.

To this end, we exploit the value of dynamic [^18^F]PI-2620 tau-PET imaging to decipher the entire A/T/N classification scheme in patients with distinct tauopathies with a single scan in a cross-sectional study design. We demonstrate that assessment of the tracer efflux via [^18^F]PI-2620_K2a_ is a highly effective kinetic PET-biomarker for distinguishing between 3/4RT and 4RT patients. Furthermore, we highlight the significance of [^18^F]PI-2620_DVR_ to quantify regional tau burden, and in conjunction with [^18^F]PI-2620_R1_, a biomarker of neuronal injury, for detailed disease staging.

We present, to the best of our knowledge, the first systematic efflux imaging data analysis of a tau-PET tracer across distinct tauopathies, both at group and individual levels. As the main finding of our study, we observed similar regional [^18^F]PI-2620 efflux for 4RT patients and healthy controls, with only minor alterations observed in the basal ganglia region, whereas, 3/4RT patients exhibited significantly reduced tracer clearance in cortical regions. This differential efflux pattern can be explained by the distinct tau isoform-specific characteristics of [^18^F]PI-2620 binding and clearance. According to previous findings, the K2a pattern, varies with tau isoform composition. In 4RT, efflux patterns resemble healthy controls due to lower binding affinity or reduced entrapment in 4R tau aggregates. In contrast, 3/4RT show higher tracer retention and reduced cortical clearance, reflecting complex binding dynamics from mixed tau isoforms [11]. Therefore, lower K2a values, align with the pathological profile characteristic of AD, highlighting that the presence of both β-amyloid and tau biomarkers, specifically 3/4R tau, is essential for a reliable AD diagnosis [23]. Notably, cortical areas exhibited the highest discriminative power, underscoring the importance of these regions in distinguishing tauopathy subtypes. Our regional analyses highlighted the high sensitivity of K2a imaging, pinpointing that reduced [^18^F]PI-2620 clearance may allow for the detection of Alzheimer’s tau pathology at lower thresholds, potentially aiding in the differentiation from other neurodegenerative disorders. When we challenged [^18^F]PI-2620 K2a in a ROC analysis against DVR, the highest discriminatory power for prediction of β-amyloid-positivity was obtained for the presented clearance biomarker. When compared against the gold standards of binary β-amyloid-PET scans and CSF Aβ_42_/Aβ_40_ ratios [28, 29], [^18^F]PI-2620 clearance showed high positive and negative predictive values, indicating its strong potential for accurate classification of the β-amyloid status. In line with these results, we were previously able to show that kinetic modeling parameters of [^18^F]PI-2620 can differentiate tau-positive clusters between 3/4R and 4R tauopathies [11].

The findings of this cohort align with several studies in which [^18^F]PI-2620 has demonstrated a higher PET signals in patients with 3/4RT compared to patients with 4RT [9, 21, 30–32], owing to its more favorable kinetic profile in the presence of 3/4R tau filaments. As the underlying mechanism of action there is evidence from i) in vitro binding assays, ii) molecular docking studies [10], and iii) autoradiography that [^18^F]PI-2620 has stronger binding affinity to 3/4R-tau compared to 4R-tau isoforms. Thus, although [^18^F]PI-2620 exhibited high affinity for both 3/4R-tau and 4R-tau isoforms in autoradiography and immunohistochemistry studies [7, 21, 33], it is reasonable that the pharmacokinetic profile of regional tracer abundance is different across distinct tauopathies. In this regard, structural biology studies using cryo-electron microscopy have already uncovered the intricate structural complexity of tau protein, demonstrating how its folding patterns and conformations vary among different tauopathies and across distinct brain regions [34–36].

Additional to different kinetic profiles, we also exploited the different topologies of tau accumulation in primary and secondary tauopathies. In line with previously published data, 4RT showed mainly subcortical tau accumulation, particularly in the globus pallidus [20, 21, 30], whereas 3/4RT patients had predominant cortical PET signals. Previous efforts to standardize quantitative tau measurements across different tracers have supported the development of a universal scale. Notable examples include CenTauR, introduced for the Alzheimer’s disease continuum [19], and histopathological defined regions for PSP as outlined by Kovacs et al [20]. As a major novelty, we provide the first proof of concept for staging distinct tauopathies within the A/T/N framework. We used K2a as a predictor for the β-amyloid status and, through region-specific separation of tau-PET binding using predefined CenTauR and Kovacs regions for AD and PSP, achieved a clear distinction between both cohorts, also accounting for neuronal injury. This ability to differentiate tauopathies in such detail lays the groundwork for a deeper understanding of the underlying mechanisms, particularly in the context of regional tau aggregation patterns.

However, when examining specific regions like the basal ganglia, we observed only a trend towards a higher [^18^F]PI-2620_DVR_ in the basal ganglia of 4RT compared to AD, which did not remain significant after multiple comparison testing. This finding may be attributed to the distinct binding characteristics of [^18^F]PI-2620 to different tau isoforms, where even a low deposition of 3/4R tau in the basal ganglia of AD (e.g., AD-CBS) could generate a higher PET signal than the substantial 4R tau deposition observed in PSP cases [21, 37]. A second ROC analysis further validated the discriminatory power of [^18^F]PI-2620 DVR with high potential to distinguish β-amyloid positivity [9]. Notably, the comparison between both ROC analyses of K2a and DVR highlighted the superior performance of K2a in discriminating β-amyloid-positivity. This finding suggests that K2a may provide a more nuanced and sensitive measure of β-amyloid-related changes, making it a valuable parameter for future research and clinical applications. Additionally, we provide evidence that [^18^F]PI-2620_DVR_ and [^18^F]PI-2620_K2a_ are strongly negatively associated in cortical regions of 3/4RT, but not in 4RT patients. The opposite was observed in the basal ganglia, highlighting the dependence of [^18^F]PI-2620 binding affinity on actual tau abundance in the target tissue. Consequently, we questioned if the solely consideration of the basal ganglia could successfully predict β-amyloid-positivity in the evaluated patients with tauopathies. The ROC analysis of K2a in the basal ganglia revealed that significant discrimination was achieved only with PET validation, however, this was not replicated with CSF or combined biomarker validation. Conversely, DVR did not demonstrate significant discriminatory power in the basal ganglia, suggesting that this region may be less effective for β-amyloid status prediction using [^18^F]PI-2620 binding.

Next, we observed that CSF p-tau-181 levels were elevated in AD compared to 4RT aligning with previous findings that associate elevated p-tau-181 levels with tau-PET specifically in AD patients [38, 39]. In contrast, 4RT patients often exhibit even lower p-tau-181 concentrations than age-matched controls, congruent with previous reports [40, 41].

As the third pillar of the proposed staging scheme, our study provides valuable insights into the utility of [^18^F]PI-2620 tracer delivery as a predictive marker for neuronal damage in patients with tauopathies. We exploited concomitant scans of FDG-PET and early-phase β-amyloid-PET, as well as CSF analysis with total-tau evaluation for gold-standard validation of early-phase tau-PET as a regional surrogate biomarker of neurodegeneration [13, 42, 43].

In line with literature, our findings showed a significant association between [^18^F]PI-2620_R1_, FDG-PET early-phase β-amyloid-PET [44]. Likewise, analysis of CSF total-tau suggested an inverse relationship between whole brain perfusion and neuronal injury [45]. We also observed a significant association between [^18^F]PI-2620_R1_ and volumetric cMRI, particularly in the 3/4RT [46] and less pronounced in the 4RT group [47, 48]. Interestingly, subcortical analysis demonstrated a divergent relationship between [^18^F]PI-2620_R1_ and atrophy in specific brain structures, consistent with previous studies reporting reduced volumes of the nucleus caudate [49] and thalamus [50, 51]. A negative correlation was found in the globus pallidus where enhanced perfusion was reported in 4RT [14] and AD [52, 53]. Reduced perfusion in the caudate nucleus, however, was associated with reduced volume in 4RT [14]. Although the mechanisms underlying these perfusion alterations are not yet fully understood, it is hypothesized that regions affected by atrophy may exhibit compensatory pathological increases in neuronal activity and neuroinflammation, resulting in elevated perfusion depending on the disease stage [54].

Finally, visual assessments of early-phase tau-PET maps revealed excellent inter-reader and inter-modality agreements. These results emphasize the reliability and consistency of visual assessments across various PET modalities and underscore the value of early-phase tau-PET imaging as a surrogate of cerebral perfusion in patients with tauopathies, with clinical implications for diagnosis and staging.

This study has several limitations, including, despite a few cases, the absence of autopsy confirmation for the clinically diagnosed cases, which could have led to diagnostic inaccuracies. Although we did not expect significant differences between PSP and CBS in the combined 4RT group, the distinct molecular mechanisms of tau pathology in these diseases could still result in varied patterns of pathological spread. Nevertheless, basal ganglia involvement remains a main characteristic feature in both PSP and CBS [30]. Importantly, the primary aim of this study was to demonstrate the feasibility of classifying individual patients within the A/T/N classification scheme using dynamic tau-PET imaging. We calculated DVR and K2a thresholds through ROC analysis, establishing cutoffs that allowed for assignment into either the β-amyloid-positive or β-amyloid-negative group. Through further quantitative regional assessment of tau load and perfusion, we were able to differentiate primary and secondary tauopathies (e.g., AD-CBS vs. 4R-CBS), which allowed us to successfully categorize and stage individuals. As a limitation, we acknowledge that our staging scheme is a preliminary approach, warranting further advancement by additional healthy controls and larger populations. Normalizing and harmonizing results across multicenter settings, similar to the Centiloid scale for β-amyloid imaging [55], will be essential. Additionally, defining cut-offs based on larger cohorts will improve the ability to discriminate between different neurodegenerative disorders. Furthermore, studies including cohorts from different centers with distinct neurodegenerative diseases like frontotemporal dementia (i.e., tau-positive non-fluent variant primary progressive aphasia) or multiple system atrophy are necessary to confirm the applicability of these findings in a real-world setting. Our results demonstrate that the accurate assessment of altered K2a is effective only in the presence of established tau pathology. Consequently, K2a is likely to lose sensitivity in the very early stages of AD. Further longitudinal studies are needed to address this gap and may capture changes in neurofibrillary profiles and the recently proposed shifts in tau isoforms throughout the trajectory of AD in vivo [56–58].

In summary, dichotomous assessment at the individual patient level revealed a strong agreement between semiquantitative and visual methods in reliably distinguishing patients with clinically diagnosed 3/4RT from those with 4RT. Importantly, this study underscores the potential of dynamic [^18^F]PI-2620 tau-PET imaging to classify and stage patients within the A/T/N framework, providing valuable insights for future clinical trials. This approach suggests that a single dynamic [^18^F]PI-2620 tau-PET scan could effectively replace redundant β-amyloid-and FDG-PET scans, thereby reducing radiation exposure for patients and lowering healthcare costs.

## Methods

### Cohort and study design

All subjects were recruited and scanned at the Ludwig-Maximilians-University of Munich (LMU), Department of Nuclear Medicine between 2018 and 2024. Patients were diagnosed to belong to the AD continuum (total n=47; ⌀75±9 years, 62% female) as 3/4RT or PSP/CBS (n=82; ⌀74±7 years, 42% female) as 4RT according to current diagnostic criteria [3, 21, 59] and compared to healthy controls (HC) (n=17; ⌀70±10 years, 53% female). Patients with AD were required to meet criteria for typical or atypical AD with MCI or dementia according to the diagnostic criteria of the National Institute on Aging and Alzheimer’s Association and were only included as AD if CSF and/or PET biomarkers of β-amyloid pathology were positive [60]. Patients with probable or possible PSP and CBS were assessed in accordance with the current diagnostic criteria, with a particular emphasis on close monitoring of disease progression during clinical follow-up (31 ± 27 months) [3, 59]. In patients with PSP, disease severity was assessed using the PSP Rating Scale, while cognitive impairment severity was evaluated using the Montreal Cognitive Assessment (MoCA). All participants (or their legal representatives) provided a written consent for PET imaging. The study protocol and PET data analyses were approved by the local ethics committee (LMU Munich, application numbers 17-569 and 19-022). The study was carried out according to the principles of the Helsinki Declaration.

### CSF analyses

A subset of the participants (3/4RT, n=41; 4RT, n=63) underwent lumbar puncture at their visit at LMU University Hospital, Munich and their CSF levels were analyzed with ELISA Innotest Kit (Fujirebio Europe N.V., Belgium) at the MVZ laboratory PD Dr. Volkmann & Kollegen GbR in Karlsruhe, Germany. The CSF biomarkers included Aβ_42_, Aβ_40_, Aβ ratio, t-tau, and p-tau-181. The respective normal cut-off values were for Aβ_42/40_ ratio>5.5%, t-tau<445pg/ml, and for p-tau-181<61 pg/ml, according to standardized laboratory diagnostics at the partnering laboratory.

### Radiosynthesis

Radiosynthesis of [^18^F]PI-2620 was achieved by nucleophilic substitution on a BOC-protected nitro precursor using an automated synthesis module (IBA, Synthera). The protecting group was cleaved under the radiolabelling conditions. The product was purified by semipreparative HPLC. Radiochemical purity was 99%. Non-decay corrected yields were about 35% with a molar activity of 8·10^6^ GBq/mmol at the end of synthesis. Radiosynthesis of Flutemetamol was performed as described previously [44, 61]. Florbetaben and FDG were purchased commercially.

### Tau-PET acquisition and preprocessing

All patients were scanned at the Department of Nuclear Medicine, LMU Munich, with a Biograph 64 or a Siemens mCT PET/CT scanner (both Siemens, Erlangen, Germany). The dynamic brain PET data were acquired in 3-dimensional list-mode over 60 minutes and reconstructed into a 336 x 336 × 109 matrix (voxel size: 1.02 x 1.02 x 2.03 mm3) using the built-in ordered subset expectation maximization (OSEM) algorithm with 4 iterations, 21 subsets and a 5 mm Gaussian filter on the Siemens Biograph and with 5 iterations, 24 subsets and a 5 mm Gaussian filter on the Siemens mCT. A low-dose CT scan preceded the PET acquisition and served for attenuation correction. Frame binning (n=35) was standardized to 12×5 seconds, 6×10 seconds, 3×20 seconds, 7×60 seconds, 4×300 seconds and 3×600 seconds.

### Spatial normalization and kinetic modelling

We acquired kinetic parameters by an inhouse automated pipeline as described previously [9, 16]. In brief, following initial motion correction of each full dynamic dataset, image-derived input functions (IDIF) were obtained through automated extraction of the PET signal from the carotid artery during the 60-minute dynamic PET scan. All images were registered to MNI space using the established [^18^F]PI-2620 PET template [62]. R1 values, indicative of delivery, and K2a efflux rate parameters, representing dissociation from the target, were extracted using simplified reference tissue modelling (SRTM2) as implemented in the Qmodelling package [63]. Distribution volume ratio (DVR) images, reflecting overall tracer binding, were calculated using the mean value from a region of interest (ROI) in the inferior cerebellar grey matter as the scaling factor. For ROI analyses, DVRs, R1, and K2a values were extracted in MNI space using delineated regions from the Brainnetome atlas [64]. To determine delivery, tracer efflux or tau-PET abnormality, z-scores were obtained for DVR, R1 and K2a values, using an in-house dataset of healthy control subjects. Static FDG-PET (30-50 min.) and early-phase β-amyloid-PET (0-10 min.) were normalized to Standardized Uptake Value ratios (SUVR), relative to the mean intensity of a whole brain VOI.

### Visual analysis of stereotactic surface projections

For visual interpretation of early-phase [^18^F]Flutemetamol_0–10min_/[^18^F]Florbetaben_0–10min_ (β-amyloid-PET_0–10min_), [^18^F]PI-2620_0.5–2.5min_ and FDG-PET images, three-dimensional stereotactic surface projections (3D-SSP) [65] were generated and assessed by three independent readers. A detailed description of the visual assessment of stereotactic surface projections is provided in the **Supplement**.

### Statistical analysis

Correlations of regional SUVR, R1, K2a, and DVR among imaging ([^18^F]PI-2620, β-amyloid, FDG, cMRI) and cerebrospinal fluid (p-tau-181, t-tau) readouts were evaluated using Pearson’s correlation coefficient (R). Quantitative variables were reported as mean ± standard deviation. An unpaired, two-tailed t-test was used for groupwise comparison of p-tau-181 and t-tau between both tauopathy cohorts. For visual analysis, the intra-reader agreement for [^18^F]PI-2620_0.5–2.5min_, β-amyloid_0-10min_ and FDG_30-50min_ was calculated using Fleiss kappa, while the intra-modality agreement was determined using Cohen’s kappa. Analysis of Variance (ANOVA) was employed including Sidak’s multiple comparisons test for DVR- and K2a z-scores of Brainnetome atlas regions.

For staging, the CenTauR mask, covering mesial temporal, meta-temporal, posterior cingulate/precuneus, and subfrontal areas, [66] was utilized in conjunction with manually delineated PSP target regions [21] in Montreal Neurological Institute (MNI) space, aligned with the tau aggregate patterns identified in histopathological studies as previously described by Kovacs et al [20]. This approach was used to generate DVR scores, ranging from 0 to 100, by referencing the cases with the lowest and highest uptake in each region across all n=129 participants, respectively. Additionally, the perfusion impairment across the combined CenTauR and Kovacs mask was assessed with the same approach, with a score of 100 representing the case exhibiting the most significant neuronal damage.

An effect size analysis of K2a and DVR scores was performed using the Brainnatome atlas, which includes 246 regions, to identify the top 10% most discriminating regions between the two tauopathy cohorts. The resulting 24 regions were then averaged and used in subsequent correlation analyses with DVR and K2a, as well as with CSF p-tau-181, and to conduct a receiver operating characteristic (ROC) analysis to predict β-amyloid positivity. ROC curves were calculated after logistic regression of K2a and DVR against β-amyloid-PET, CSF and both biomarkers combined. The optimal cutoff for β-amyloid positivity using K2a was estimated using the Youden index. Comparison of the area under the curve (AUC) of ROC curves between K2a and DVR were conducted using a nonparametric approach as previously described by DeLong et al. [67]. A significance level of p < 0.05 was applied in all analyses. All statistical analyses were performed using SPSS (version 27.0, IBM, New York, USA) and Graph Pad Prism (V9, GraphPad Software).

## Data Availability

The datasets generated during and/or analyzed during the current study are available from the corresponding author on reasonable request.

