## Supplemental Material for "Assessment and staging of A/T/N with a single dynamic [^18^F]PI-2620 recording"

**Supplemental Methods**

*Visual analysis of stereotactic surface projections*

For visual interpretation of early-phase [^18^F]Flutemetamol_0–10min_/[^18^F]Florbetaben_0–10min_ (β-amyloid-PET_0–10min_), [^18^F]PI-2620_0.5–2.5min_ and FDG-PET images, three-dimensional stereotactic surface projections (3D-SSP) [26] were generated using the software Neurostat (Department of Radiology, University of Washington, Seattle, WA, USA). Voxel-wise Z-scores were calculated in Neurostat by comparing the individual tracer uptake of β-amyloid-PET_0–10min_, [^18^F]PI-2620_0.5–2.5min_ and FDG to a historical FDG-PET database (cognitively healthy individuals, age-matched, N = 18). Three experienced nuclear medicine physicians (J.G., P.B., and M.B.) visually assessed the 3D-SSP images using the Z-score maps (GBM scaling) and rated the severity of perfusion and metabolic reduction in cortical regions used in the clinical routine (frontal, parietal, temporal, occipital cortex areas) from 0 to 3 (0 = no reduction, 1 = mild, 2 = intermediate, 3 = severe). Based on the individual readers assessment, whole-brain 3D-SSP images were rated binary (0/1) and according to the severity of present neurodegeneration (0 = no/1 = mild/2 = intermediate/3 = severe neurodegeneration). Readers had access to the pre-PET clinical diagnosis and to the scan modality. Prior to the visual read, all three readers received detailed instructions including 10 training sets. Reading was performed in balanced order to avoid systematic bias. Reader 1 started with FDG-PET, continued with β-amyloid-PET and [^18^F]PI-2620_0.5–2.5min_; Reader 2 started with [^18^F]PI-2620_0.5–2.5min_, continued with FDG-PET and β-amyloid-PET_0–10min_; Reader 3 started with β-amyloid-PET_0–10min_, continued with [^18^F]PI-2620_0.5–2.5min_ and finished with FDG-PET. Overall, n=217 reads were carried out by each reader for 3/4R-Tauopathies (n=77; [^18^F]PI-2620_0.5–2.5min_: n=38, β-amyloid-PET_0–10min_: n=24 and FDG-PET: n=15) and 4R-Tauopathies (n=140; [^18^F]PI-2620_0.5–2.5min_: n=68, β-amyloid-PET_0–10min_: n=36 and FDG-PET: n=36). Due to artifacts that occurred during normalization to the 3D-SSP (e.g., dilatation of ventricular system, pronounced atrophy, post-resection), one FDG-PET scan was excluded from the 3/4RT cohort for further analysis. In the 4RT group, two FDG-PET scans, two early-phase β-amyloid-PET scans, and two [^18^F]PI-2620 scans were excluded.

*AI Algorithm for volumetric MRI*

For quantitative analysis of brain MRI, the commercially available AI tool "mdbrain", version 3.3.0 (mediaire GmbH, Berlin) was used. The system leverages a custom deep learning segmentation model based on the U-Net architecture [63] to perform a highly accurate hemisphere- and region-specific (e.g., lobes) rapid brain volumetry in clinical routine settings, which was trained on a heterogeneous dataset of 3D T1w images (n = 1,851, balanced male/female). Augmentation techniques (augmentation of contrast, resolution, rotation, elastic deformation) have been used to maximize the model’s applicability in daily routine. The volumes of 18 brain regions including the hippocampus are determined and percentiles are derived by comparison to a cohort of healthy individuals (n = 3,179, balanced m/f, age range 18 to 92), while accounting for age, sex, and total intracranial volume.

**Supplemental Results**

*Delivery of [^18^F]PI-2620 depicts neuronal damage in patients with tauopathies (N)*

We performed an intermodal correlation analysis between [^18^F]PI-2620_R1_, FDG-PET and early-phase β-amyloid-PET in our combined tauopathy cohort. We observed a significant correlation between [^18^F]PI-2620_R1_ and FDG-PET in the frontal cortex (R=0.24; P=0.017), the temporal cortex (R=0.26; P=0.0089), the parietal cortex (R=0.53; P<0.0001), the occipital cortex (R=0.31, P=0.0019), and the basal ganglia (R=0.24; P=0.0018). The insular cortex, although indicating a clear trend, did not fully reach the significance threshold of P<0.05 (R=0.19; P=0.065). Similarly, [^18^F]PI-2620_R1_ was significantly associated with early phase β-amyloid-PET in the frontal cortex (R=0.21; P=0.023), the temporal cortex (R=0.38, P<0.0001), the parietal cortex (R=0.56; P<0.0001), the insular cortex (R=0.32; P=0.0003), the occipital cortex (R=0.42; P<0.0001) and the basal ganglia (R=0.29; P=0.0011) (**Supplemental Figure 1**).

Additionally, in a subset of 4RT (n=43) and 3/4RT (n=22), volumetric MRI was acquired to obtain atrophy scores (in %) for both cortical hemispheres and hindbrain regions. Subcortical bilateral volumes were successfully obtained in 4RT (n=32) and 3/4RT (n=12), respectively (**Supplemental Figure 3**). When correlating [^18^F]PI-2620_R1_ with the degree of atrophy, a significant positive association was observed in all cortical regions of 3/4RT including the frontal cortex (R=0.57; P<0.0001), the temporal cortex (R=0.41; P=0.0051), the parietal cortex (R=0.64; P<0.0001) and the occipital cortex (R=0.49; P=0.0007). Perfusion deficits in the cortex of 4RT were significantly correlated with atrophy in the parietal lobe (R=0.23; P=0.03), while in the frontal lobe, only a positive trend was indicated (R=0.20; P=0.065). No significance, however, was attained in the temporal (R=0.07; P=0.50) and occipital cortex (R=0.07; P=0.51) (**Supplemental Figure 3A**). Considering the subcortical regions, [^18^F]PI-2620_R1_ was negatively correlated with atrophy in the globus pallidus (R=-0.29; P=0.018) of 4RT), while a moderate degree of correlation was found in the nucleus caudatus (R=0.27; P=0.03). The relative perfusion of the putamen of the same patient sample was borderline negatively associated with atrophy (R=-0.24; P=0.053) among both imaging modalities. In the 3/4RT group, only the putamen revealed a significant negative association among [^18^F]PI-2620_R1_ and the level of atrophy (R=-0.44; P=0.031) (**Supplemental Figure 3B**).

**Supplemental Figure 1**

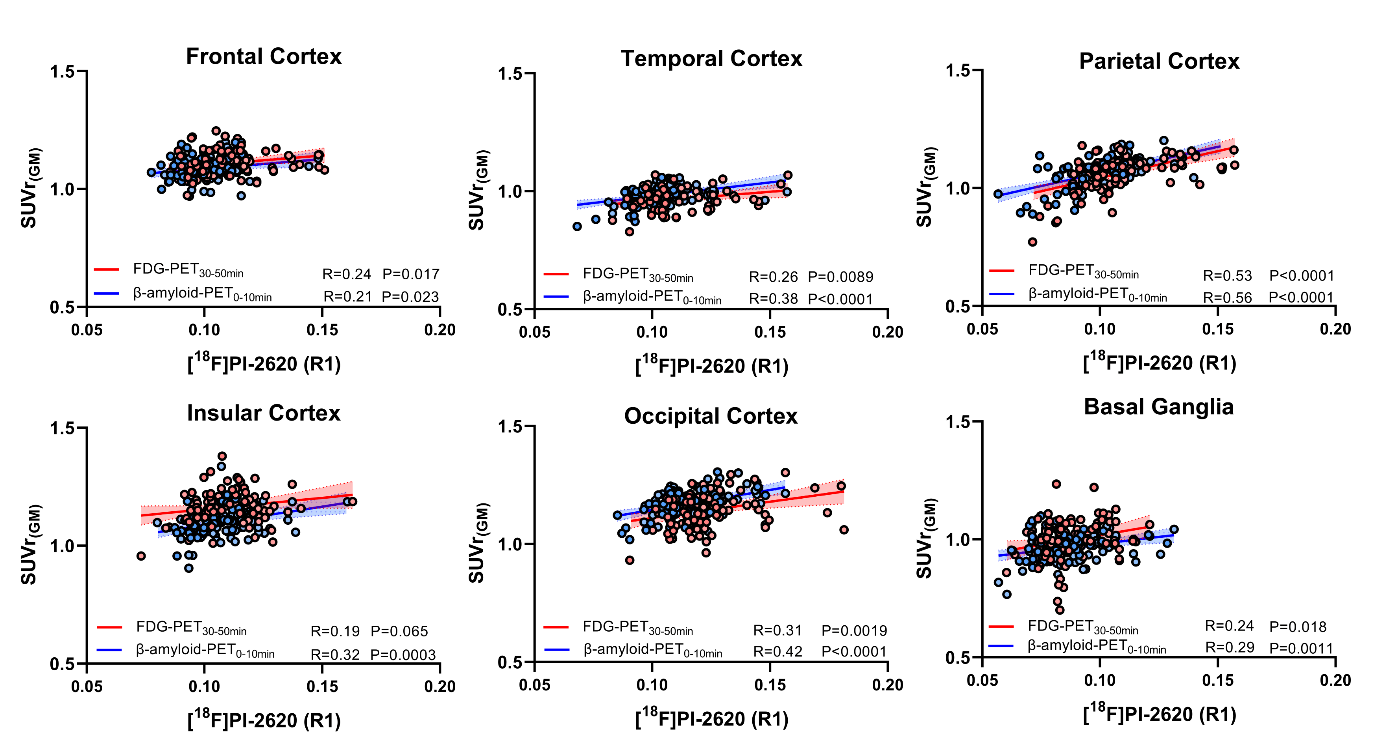

**Supplemental Figure 1: Relationship between the relative perfusion (R1) of [^18^F]PI-2620 and early-phase β-Amyloid-PET, FDG-PET.** Whole brain scaled SUVR of early phase β-amyloid-PET (blue dots) and FDG-PET (red dots) are correlated with [^18^F]PI-2620_R1_ in cortical and subcortical regions of 4RT and 3/4RT.

**Supplemental Figure 2**
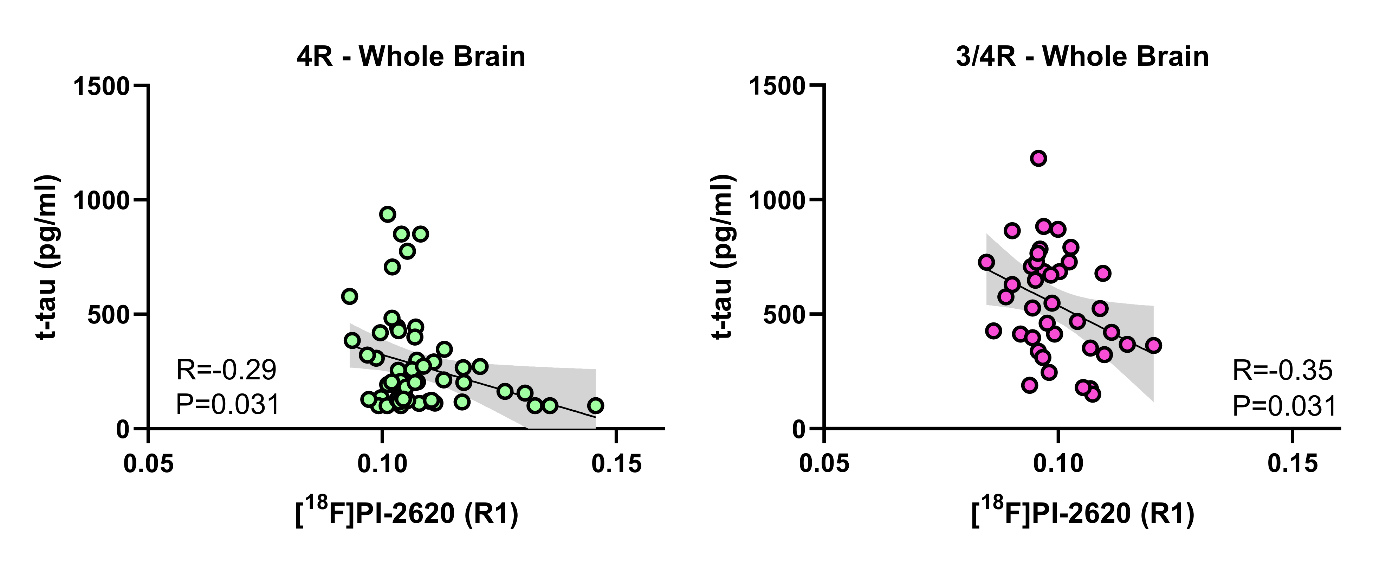
**Supplemental Figure 2: Relationship between the relative perfusion (R1) of [^18^F]PI-2620 and total tau protein.** Green and purple scatterplots depict negative agreement among whole brain [^18^F]PI-2620_R1_ and total tau protein in the cerebrospinal fluid of both cohorts.

**Supplemental Figure 3**

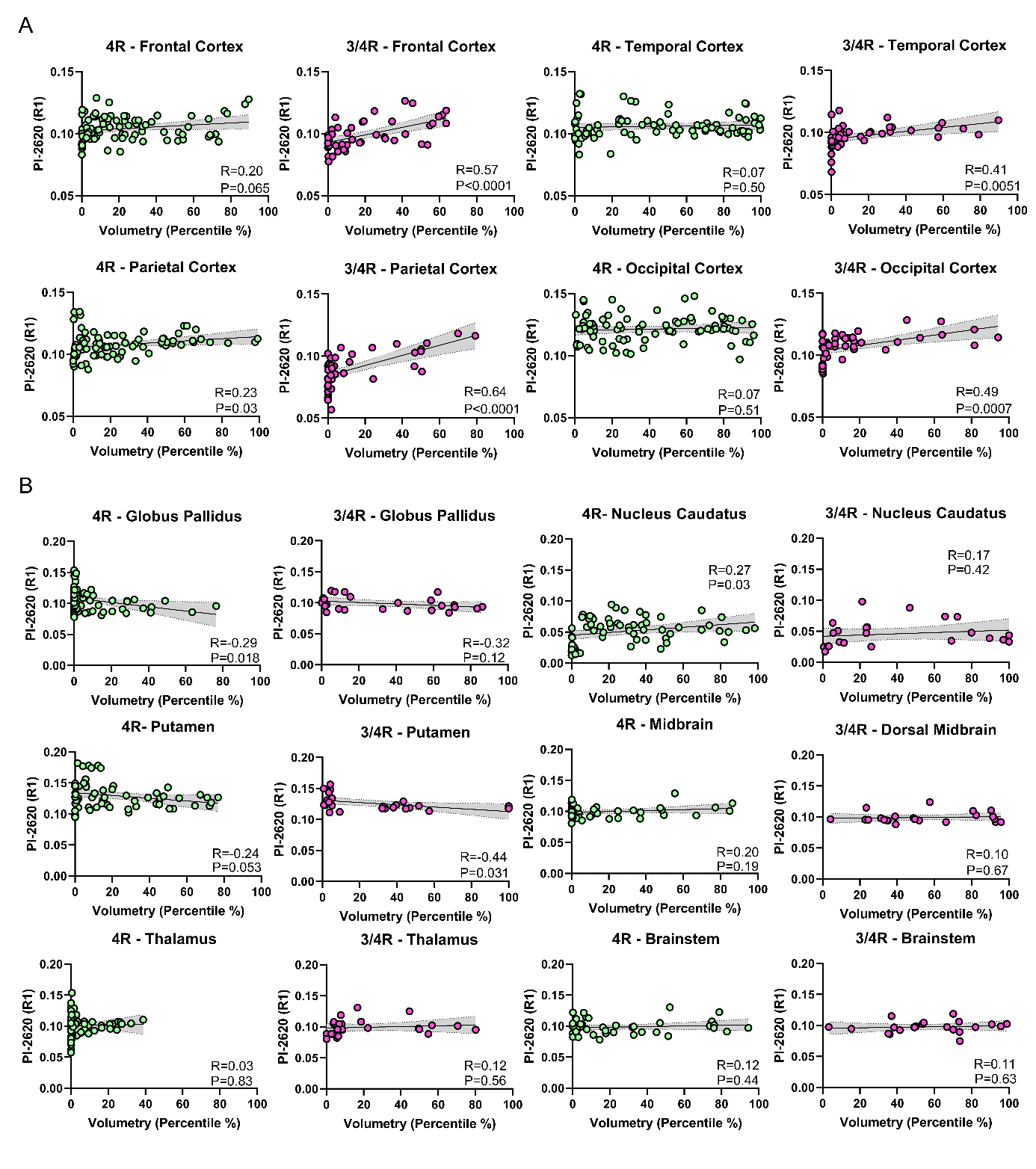
**Supplemental Figure 3: Correlation charts of [^18^F]PI-2620_R1_ and atrophy in distinct brain regions.** (**A-B**) Correlation plots depict the agreement of [^18^F]PI-2620_R1_ and volumetric MRI-acquired atrophy in cortical (**A**) and subcortical (**B**) brain regions of 4RT (green dots) and 3/4RT (purple dots) participants.

**Supplemental Figure 4**

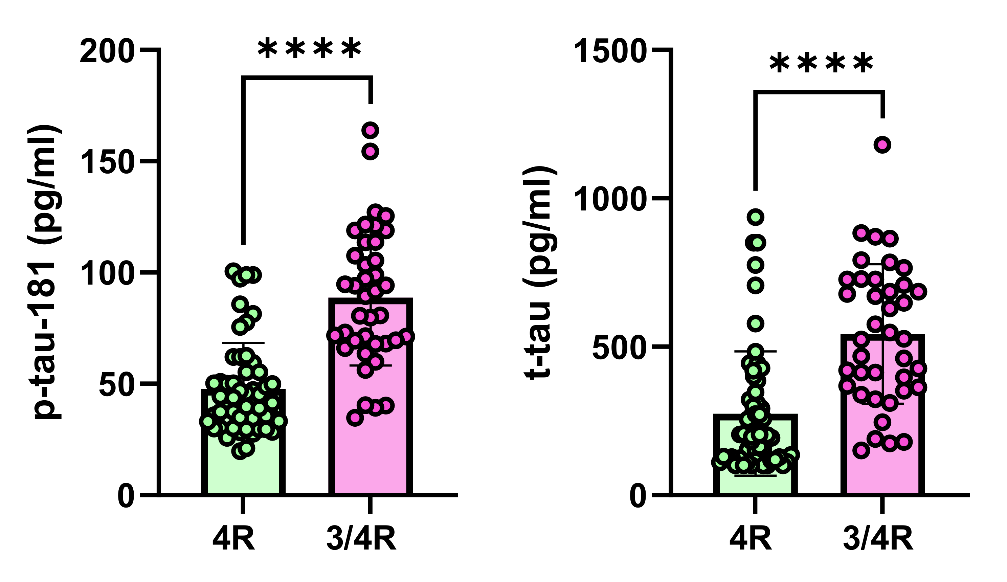

**Supplemental Figure 4: Groupwise comparison of p-tau-181 and t-tau.** 4RT (n=56) and 3/4RT (n=39) were compared using an unpaired, two-tailed t-test.
